# The role of school reopening in the spread of COVID-19

**DOI:** 10.1101/2020.09.03.20187757

**Authors:** Richard Beesley

## Abstract

Many countries chose to close schools as part of their response to the SARS-CoV2 coronavirus (COVID-19) pandemic. Whilst nations are gradually reopening schools, and many politicians advise that schools remain safe and the risks of increases in the spread of COVID-19 are low, little evidence has been presented to confirm those statements.

A review of the numbers of new confirmed COVID-19 cases by country suggests that the reopening of schools is likely to be a driver in the increase of the number of new cases. This is likely exacerbated by accompanying changes and easing of restrictions. However, with the exception of China, notable for its robust test, track, trace, and isolate processes, no other countries that had significant numbers of COVID-19 cases have successfully reopened schools without an increase in cases as a consequence.

Whilst reopening of schools following an initial peak and decrease in COVID-19 infections is desirable for a range of reasons, doing so without adequate controls and protections may lead to an exacerbation of spread within the school environment, which could then lead to increased community spread of disease.

## Introduction

2 September 2020.

To date, the SARS-Cov2 coronavirus infection (COVID-19) has infected over 26 million individuals worldwide, and been responsible for over 860,000 deaths globally. Whilst our understanding of the route for spread of the virus has developed, the role of specific aspects of our communities is not yet fully understood. Many countries closed schools and colleges early in the outbreak of COVID-19 in an effort to reduce the spread. Some have since re-opened schools, although this may have contributed to an increase in the number of confirmed infections. Many governments have claimed that school reopening either has not, or will not, lead to an increase in COVID-19 cases, yet have produced little evidence to support that claim.

This paper is based on a review of publicly available national data from countries that have experienced substantial numbers of COVID-19 cases to assess the potential role of reopening schools in the spread of the virus and increases in numbers of cases.

## Methods

The number of new daily confirmed cases of COVID-19 per country^1^ was downloaded, up to 1 September 2020. From this, seven-day rolling totals were calculated for each country by day. This process removed variability in the data that was due to some testing or analysis centres not operating at full capacity every day of the week, which is the case in some countries. The difference in monitoring and tracking between countries is noted, although data from each country is analysed separately so these differences are less critical.

The final analysis includes countries that experienced a seven-day total of at least 5,000 new cases on at least one day since they started recording COVID-19 cases, where there is at least one clear wave or peak in cases (whether this has been followed by a second peak or not). This cut-off was selected because small numbers of cases can easily adversely affect and mask overall trends.

Countries were excluded if the initial wave of COVID-19 cases remains ongoing, if the data was highly variable (suggesting inconsistencies in case or data management), or if disparate schooling systems and dates are used throughout the country making analysis of national data meaningless (such as the USA).

For each country included, an initial analysis of the seven-day rolling total number of new cases was plotted over time. Alongside this we reviewed information about school closures due to COVID-19, school reopening, and school holiday dates. This data was acquired from national education departments and mainstream media; where media articles were used to identify school closure and reopening dates they were verified using multiple independent sources.

Since re-opening schools, some countries have closed individual schools or those in specific areas to control the spread of the virus. These cases are not included in the analysis presented here.

For clarity, reopening of schools for the purposes of this analysis includes only in-person teaching on-site. Remote, distant and web-based teaching is excluded because, by their very nature, they do not increase the risk of transmission of COVID-19.

## Findings

Our analysis identifies 4 main patterns in which the policy decision and timing of school closures and reopening is mapped onto the total number of new cases, and two potential impacts – cases increase, or cases do not increase (Figure 1**Error! Reference source not found.**).

A. Schools have closed and not reopened. The total number of cases is therefore not related to school openings. These countries can be further divided into two – those where the number of cases has started to rise again (A1), and those where the number of cases has remained low (A2).
B. Schools reopened before or during an initial or ongoing increase in cases.
C. Schools reopened after an initial wave of cases had reduced. Again, these countries can be further divided into two – those where the number of cases has started to rise again (C1), and those where the number of cases has remained low (C2).
D. Schools did not close.

**Figure 1.**
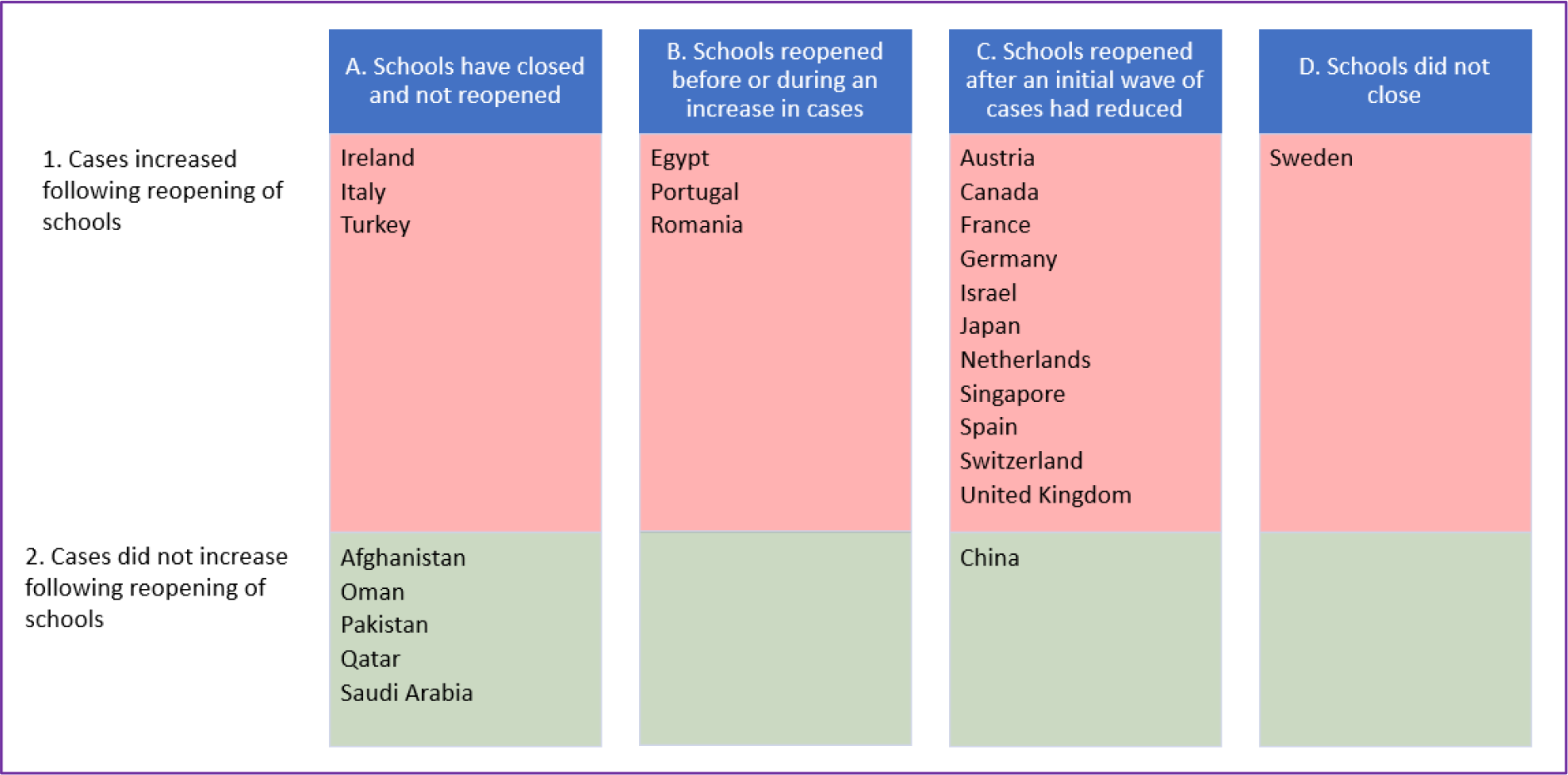
Categories of the national decisions about school reopening and the impact on the number of new confirmed cases of COVID-19 (based on 7-day rolling totals).

In countries where schools were closed and have not yet reopened, there are two distinct patterns.

Three countries (Ireland, Italy and Turkey) have seen an increase in the number of cases despite not reopening schools (Figure 2). These examples highlight that increases cannot exclusively be down to schools reopening, but due to a more complex mix of societal factors. Lifting of restrictions on groups of people meeting and travelling, and an increase in holidays (both within the country and with visitors from elsewhere) may have contributed to the rise seen.

**Figure 2.**
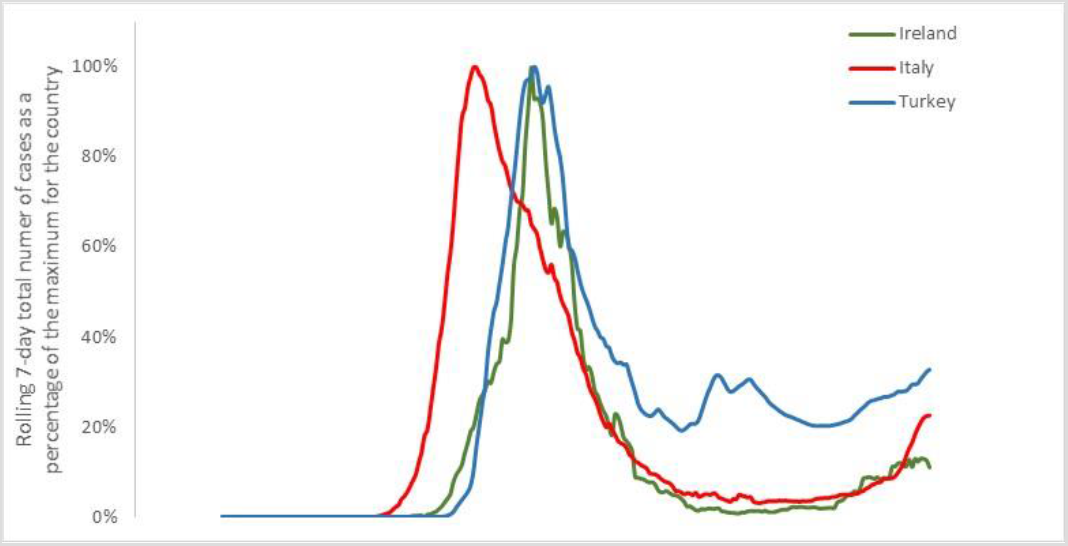
The trend of cases in three countries that closed schools but have not yet reopened them, yet have seen an increase in cases. Line is shown as a percentage of the maximum for each country to give comparability, based on the 7-day rolling total number of new confirmed COVID-19 cases.

In contrast five countries that have not reopened schools have not seen an increase in cases (Figure 3). This may be due to reduced inter-country tourism, coupled with a less prominent easing of restrictions.

**Figure 3.**
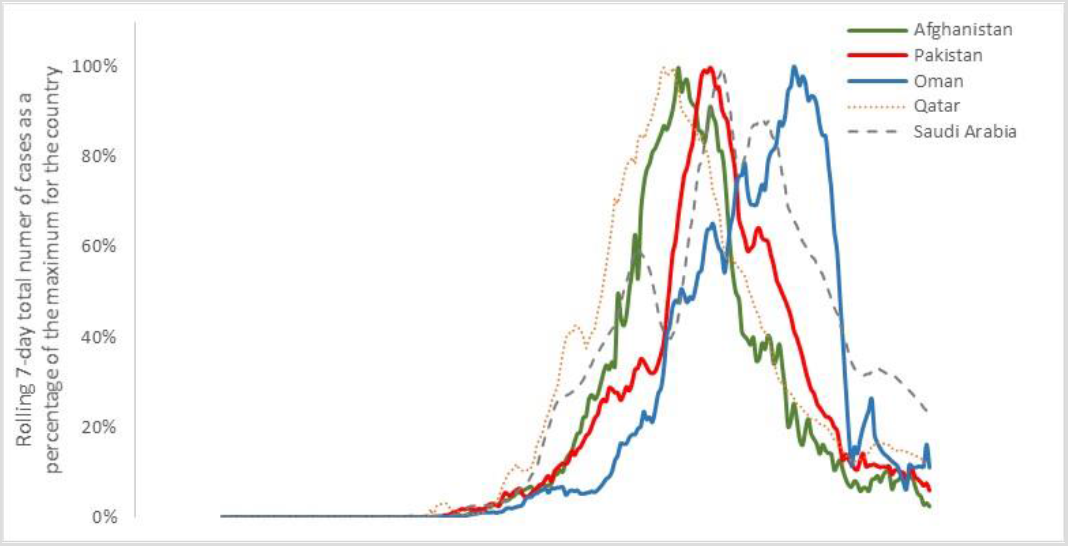
The trend of cases in five countries that closed schools but have not yet reopened them, and have not seen an increase in cases. Line is shown as a percentage of the maximum for each country to give comparability, based on the 7-day rolling total number of new confirmed COVID-19 cases.

One country (Egypt) within the analysis reopened schools before the initial wave of cases had peaked; two others (Portugal and Romania) reopened schools whilst a subsequent increase in cases was ongoing. As a result, any impact of school reopening is masked by the overall increase in cases (Figure 4).

**Figure 4.**
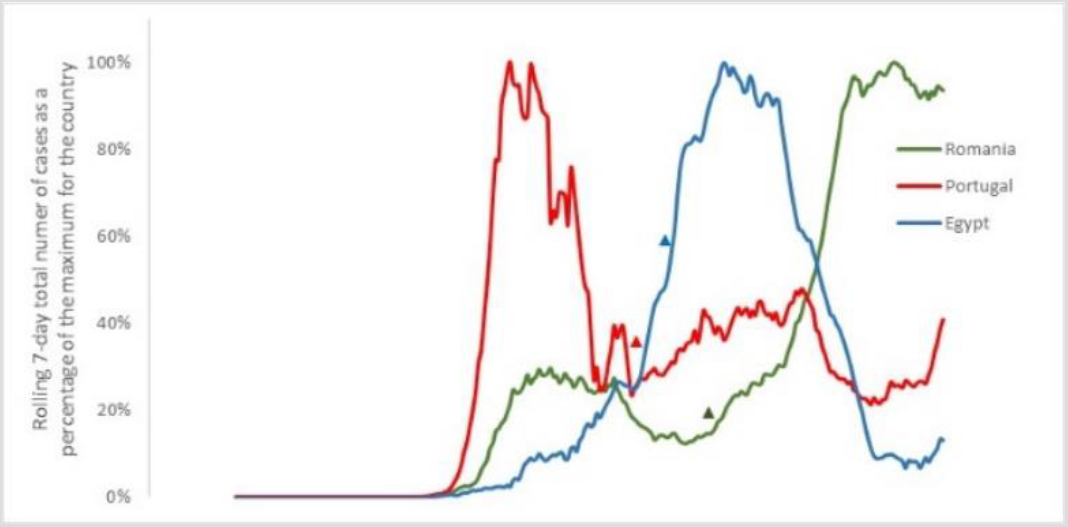
The trend of cases in three countries that closed schools and reopened them when cases were already increasing. Line is shown as a percentage of the maximum for each country to give comparability, based on the 7-day rolling total number of new confirmed COVID-19 cases. Date of school reopening is indicated by a triangle.

The majority of countries within the analysis have reopened schools following an initial peak in cases. Only one of these (China) has seen a sustained low number of cases (Figure 5). This is likely due to their robust and rapid response to suspected and confirmed cases of COVID-19, including widescale testing within the community, strict isolation measures, and immediate closure of schools with a potential outbreak.

**Figure 5.**
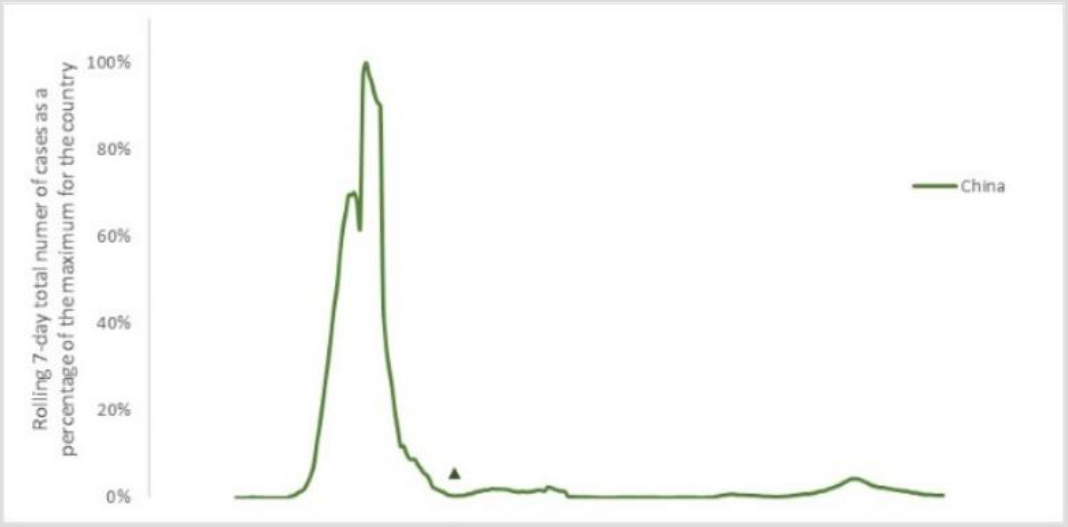
The trend of cases in China, which reopened schools and saw no increase in cases of COVID-19 as a result. Line is shown as a percentage of the maximum for each country to give comparability, based on the 7-day rolling total number of new confirmed COVID-19 cases. Date of school reopening is indicated by a triangle.

Most countries that have reopened schools after an initial peak have seen a further rise in cases. Of these, the number of days between schools reopening and the start of the increase in cases ranged from 24 to 47 days, with a mean of 36·5 days (Figure 6 and Table 1).

**Figure 6.**
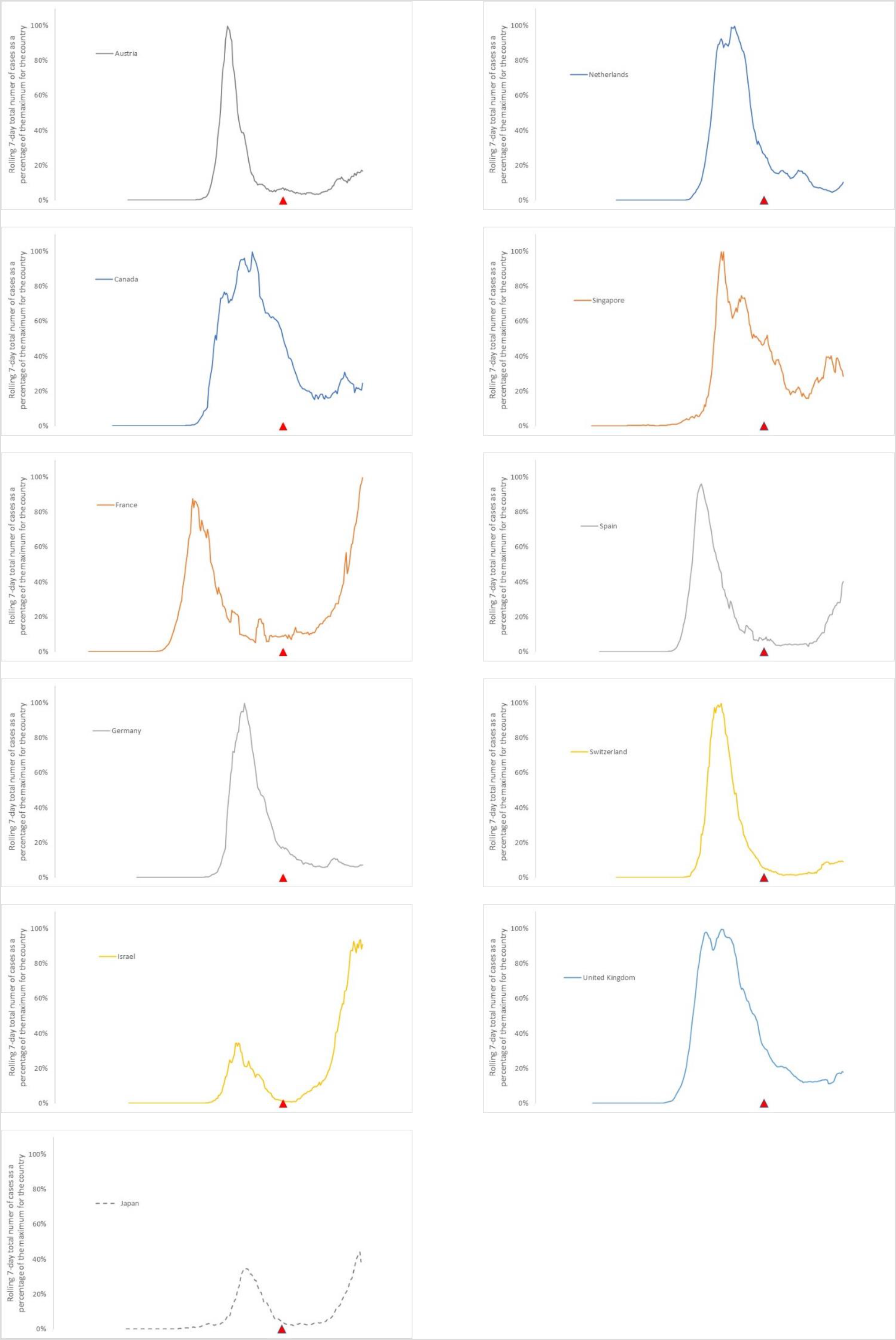
Trends of new cases in each of the countries that reopened schools and saw an increase in cases of COVID-19 as a result. Lines shown as a percentage of the maximum for each country to give comparability, based on the 7-day rolling total number of new confirmed COVID-19 cases. Date of school reopening is indicated by a triangle. For comparability, all school reopening dates have been aligned vertically.

**Table 1.** The time in days between school reopening and the start of an increase in new cases that followed, based on rolling 7-day total number of new confirmed cases of COVID-19. The Netherlands reopened primary schools before secondary schools, and experienced a peak 24 days after opening primary schools (*), and a more noticeable peak 40 days after opening secondary schools (**).

| Country | Days | Note |
| --- | --- | --- |
| Austria | 29 days | Exacerbated during summer holidays. |
| Canada | 47 days | School reopening was limited, partial and geographically restricted. |
| France | 28 days | Step change in increased cases. Exacerbated during summer holidays. |
| Germany | 37 days | Schools closed for summer holiday ahead of ongoing peak. |
| Israel | 35 days | Earlier recognised outbreaks in schools (10 days after opening). |
| Japan | 28 days | Schools reopened partially, and over a period of time. |
| Netherlands | 24 days*<br>40 days** | The peak 24 days after primary schools reopened was small but noticeable. The second, after secondary schools reopened, was much larger. |
| Singapore | 42 days | Controls introduced in schools. |
| Spain | 44 days | School reopening was limited. |
| Switzerland | 47 days | Cases continue to rise after schools close. |
| United Kingdom | 38 days | Partial school reopening. |

One country adopted a fundamentally different approach. Sweden chose not to close schools during their initial outbreak, and politicians from that country have announced that keeping schools open did not lead to an increase in cases. However, it is notable that schools in Sweden closed for the summer on 9 June; the reduction in cases (based on the seven-day rolling total) started to reduced 23 days later (Table 2 and Figure 7). This timescale is within the range identified for increases in cases after schools did reopen in other countries. As a result, it is arguable that keeping schools open may have contributed to an ongoing spread of the disease.

**Table 2.** The time in days between school closure and the start of a decrease in new cases that followed in Sweden, based on rolling 7-day total number of new confirmed cases of COVID-19.

| Country | Days | Note |
| --- | --- | --- |
| Sweden | – 23 days | Cases reduced 23 days after schools closed |

**Figure 7.**
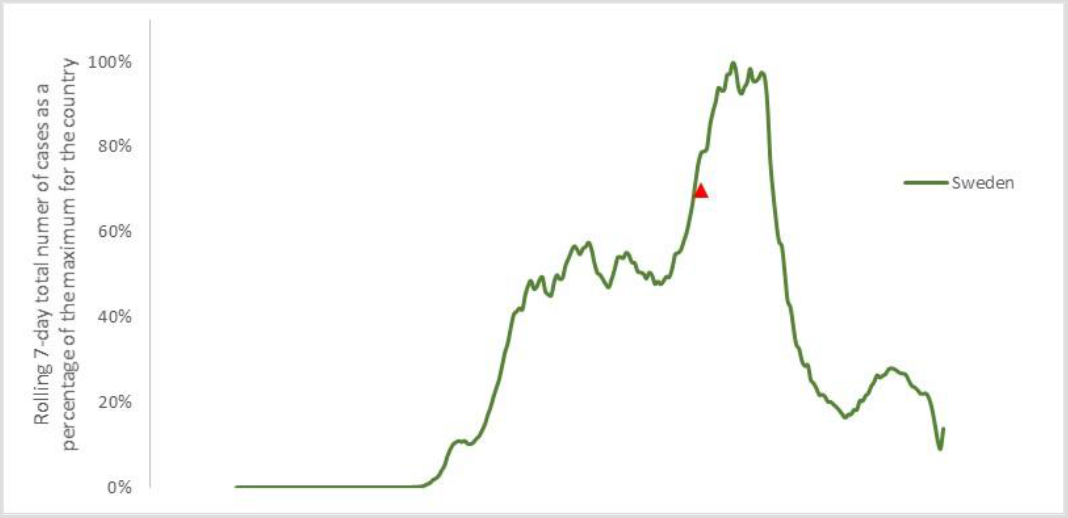
Trends of new cases in Sweden, which closed schools for the summer holiday and saw a decrease in cases of COVID-19 as a result. Line shown as a percentage of the maximum for each country to give comparability, based on the 7-day rolling total number of new confirmed COVID-19 cases. Date of schools closing is indicated by a triangle.

## Interpretations

In many countries that closed schools during the initial increase in cases of COVID-19, reopening after the number of cases reduced is associated with a new increase in cases. The different patterns seen in case loads matched against school reopening dates and processes indicate a number of contributory causes. However, the spread of the virus through schools and then into the community does appear to contribute to overall numbers of cases.

Where schools have not yet reopened, the indication is that the spread of the virus depends on a number of other factors. We hypothesise that these include the lifting of other restrictions and increased internal and into-country tourism. Thus, where these changes have taken place and tourism and travel have increased (including from areas and countries with higher underlying rates of COVID-19 infection), local numbers have started to increase. This is seen in Ireland, Italy and Turkey. By comparison, countries where schools have remained closed but other restrictions and inter-country travel remains limited, cases have not increased.

Countries which have reopened schools, either in full or in part, give a useful insight into the role schools may play in spreading the virus. The increases seen are greater than would be expected from the lifting of travel restrictions alone. Given the narrow timeframe in which cases rose after schools reopened, it is indicative that the reopening of schools contributed to the increase in cases. Further, countries that had more limited opening (Canada, Spain, United Kingdom) had a longer lead-in before the increase in cases. Similarly, Singapore, which introduced substantial controls to limit the spread of the virus within school communities, had a longer lead-in time (42 days) than other countries. Those countries that opened schools fully with limited controls saw a shorter lead-in time before an increase in cases.

Sweden is unique, in that it did not close schools. However, they are a strong example of the impact of closing schools, with a substantial reduction in cases starting 23 days after schools closed for the summer holiday period.

## Discussion

Schools form one part of our dynamic, diverse and complex communities. It is not possible to entirely separate the role of schools in the spread of the virus from other factors, notably because reopening of schools leads to parents returning to workplaces, other family members taking on care responsibilities (such as grandparents), and is often accompanied by easing of other restrictions (including travel, shops and leisure activities). It is therefore very difficult to specify with certainty that school reopening alone is responsible for an increase in the number of COVID-19 cases within a community.

However, given each of the countries that has reopened schools within this research have seen an increase in cases (with the particular exception of China), and this increase is greater than that seen in countries that have kept schools closed, it is apparent that school reopening and the associated changes in society and community transmission does play a part in COVID-19 spread.

In addition, the onset of summer holidays has exacerbated the spread of the virus, masking in part the direct effects of schools. However, school reopening does seem to have an impact. In Scotland and Northern Ireland, schools reopened around 2 weeks before England. Within those two weeks, 53 schools had reported outbreaks, with 106 students and teachers infected, and 19,000 in quarantine.

In the UK, adequate spacing and ventilation within classrooms is largely impossible. Masks are optional in many schools, and not permitted in others or in certain situations. Published risk assessments and controls within schools focus on fomite rather than droplet transmission, largely ignoring aerosol transmission of the virus, and relying on symptoms to isolate infected individuals. Given many carriers of the virus do so asymptomatically, this approach appears futile. As a result, it seems likely that an increase in viral cases will become apparent within the coming weeks in the UK.

China is an example of effective use of controls, track, trace and isolation to restrict the spread of the virus. Without those controls, the spread of the virus appears inevitable, exacerbated by school opening and travel of infected people.

## Data Availability

All data is publically available.

## Footnotes

1 Max Roser, Hannah Ritchie, Esteban Ortiz-Ospina and Joe Hasell (2020) – “Coronavirus Pandemic (COVID-19)”. Published online at OurWorldInData.org. Retrieved from: ‘https://ourworldindata.org/coronavirus’ [Online Resource]

## Notes

### Competing Interest Statement

The authors have declared no competing interest.

### Funding Statement

This work was unfunded.

### Author Declarations

Exempt from IRB / oversight.

### Summary of Updates

Updated contact details.

